# Monitoring and understanding household clustering of SARS-CoV-2 cases using surveillance data in Fulton County, Georgia

**DOI:** 10.1101/2022.03.02.22271814

**Authors:** Carol Y. Liu, Sasha Smith, Allison T. Chamberlain, Neel R. Gandhi, Fazle Khan, Steve Williams, Sarita Shah

## Abstract

**Background:** Households are important for SARS-CoV-2 transmission due to high intensity exposure in enclosed living spaces over prolonged durations. Using contact tracing, the secondary attack rate in households is estimated at 18-20%, yet no studies have examined COVID-19 clustering within households to inform testing and prevention strategies. We sought to quantify and characterize household clustering of COVID-19 cases in Fulton County, Georgia and further explore age-specific patterns in household clusters.

**Methods:** We used state surveillance data to identify all PCR- or antigen-confirmed cases of COVID-19 in Fulton County, Georgia. Household clustered cases were defined as cases with matching residential address with positive sample collection dates within 28 days of one another. We described proportion of COVID-19 cases that were clustered, stratified by age and over time and explored trends in age of first diagnosed case within clusters and age patterns between first diagnosed case and subsequent household cases.

**Results:** Between 6/1/20–10/31/21, there were 106,233 COVID-19 cases with available address reported in Fulton County. Of these, 31,449 (37%) were from 12,955 household clusters. Children were more likely to be in household clusters than any other age group and children increasingly accounted for the first diagnosed household case, rising from 11% in February 2021 to a high of 31% in August 2021. Bubble plot density of age of first diagnosed case and subsequent household cases mirror age-specific patterns in household social mixing.

**Discussion:** One-third of COVID-19 cases in Fulton County were part of a household cluster. High proportion of children in household clusters reflects higher probability of living in larger homes with caregivers or other children. Increasing probability of children as the first diagnosed case coincide with temporal trends in vaccine roll-out among the elderly in March 2021 and the return to in-person schooling for the Fall 2021 semester. While vaccination remains the most effective intervention at reducing household clustering, other household-level interventions should also be emphasized such as timely testing for household members to prevent ongoing transmission.

## Background

Understanding the spread of SARS-CoV-2 has been of critical importance^1–5^. From the perspective of public health policy and practice, identifying high-risk settings where COVID-19 transmission occurs provides important insights for targeting interventions such as contact tracing and directed testing efforts to reduce further disease spread. Despite intense scrutiny and high public interest in large superspreader events ^6,7^, smaller clusters of household cases have collectively more impact on total case counts. For example, investigations of the initial SARS-CoV-2 outbreak in Wuhan, China found that 78-85% of infection clusters occurred in families^8^, a trend that continued even after relaxation of the most stringent lockdown measures that confined cases and their contacts at home^9^.

Household contacts are particularly vulnerable to SARS-CoV-2 transmission. Household members experience high intensity exposure over prolonged durations and share enclosed and, at times, crowded living environments^10^ – factors that together increase the probability of transmission from an infected individual to a susceptible household contact^11–13^. While non-pharmaceutical interventions such as mask-wearing and social distancing effectively reduce community transmission of SARS-CoV-2^14,15^, consistently adopting such stringent preventative measures is difficult in practice, particularly in a household. Furthermore, cases can be infectious prior to symptom-onset^16^, which precludes index household cases from taking early preventative measures to protect household members. Several systematic reviews drawing data from multiple countries highlight the significance of household transmission in sustaining the COVID-19 pandemic. These reviews estimate the household secondary attack rate of the original variant to be between 16.4–30%^17–21^ and higher based on preliminary evidence for alpha (24.5% from meta-analysis of three studies^22^) and delta variants^23,24^. Finally, the household presents a unique social context where intergenerational contact between children, parents and grandparents is higher than other social settings, such as work and school; in those settings, individuals tend to be in contact with other individuals of similar age^25–29^. As such, households can be an important setting for transmission from children to older adults who have increased susceptibility^30^ and heightened probability of severe disease^31^.

Despite known heightened transmission between household members and the importance of households in the context of intergenerational transmission, there is limited quantification of the extent of household clustering of COVID-19 in the US. We sought to use surveillance data to quantify the extent of household clustering of COVID-19 among confirmed COVID-19 cases in Fulton County, Georgia. Fulton County, located in metropolitan Atlanta, is an urban county in Georgia with a population of 1.1 million. We postulate that household clustered cases continue to account for a substantial proportion of cases detected by routine surveillance. We further explore temporal trends in clustering, the distribution of household-clustered cases among key demographic groups and focus our analysis on age profiles of cases in household clusters, exploring trends in age of first diagnosed case within clusters and age patterns between first diagnosed case and subsequent household cases. Our analysis uniquely leverages a robust and large public health database of routinely-collected COVID-19 case data to identify temporally clustered cases residing at the same residential address and quantify household clustering behavior.

## Methods

### Study design and population

We conducted a retrospective cohort analysis of persons residing in Fulton County, Georgia, who were diagnosed with PCR-confirmed SARS-CoV-2 infection. We obtained data for COVID-19 cases from Georgia Department of Public Health’s (DPH) State Electronic Notifiable Disease Surveillance System (SENDSS) for Fulton County between June 1^st^, 2020 and Oct 31^st^, 2021. Per the state’s COVID-19 response statutes, all individuals with a positive diagnosis for SARS-CoV-2 must be notified to the Georgia DPH. All reported cases are captured by DPH into the SENDSS database. Cases prior to June 1^st^, 2020 were excluded due to the limited availability of testing early in the pandemic where only those with selected risk factors (e.g., age), COVID-related symptoms, or known exposure were eligible for testing. These eligibility criteria were removed in Fulton County in early June, 2020, such that all persons could access free testing, regardless of symptoms or risk factors. Cases after Oct 31^st^, 2021 were excluded as cluster data are likely incomplete due to ongoing household transmission chains.

### Definitions and outcome measures

To identify cases originating from the same residence, we standardized addresses using a geocoder which cross-referenced the case addresses with the US Postal Service address database and only cases with a valid and complete residential address were retained. Since our primary interest was in case clustering at residential locations, addresses belonging to communal residential locations (e.g., nursing homes, correctional facilities, homeless shelters and dormitories) were excluded, as were apartment addresses missing the unit number where cases from multiple units could be erroneously classified as from the same household.

Clustered cases were defined as cases with a perfectly matching standardized street address, including unit number for apartment complex addresses. Household clusters were defined as >2 COVID-19 cases residing at the same residential address with positive sample collection dates within 28 days of one another (Supplementary Figure 1). With a median incubation period estimated at 5.1 days^32^ and a median infectious period estimated at between 7-10 days^33^, the 28 days would cover two infectious periods, one incubation period, and a 3-4 day lag between symptom-onset and positive sample collection^34^. Given the high proportion of asymptomatic and undiagnosed COVID-19 cases, this would allow two diagnosed cases with one undiagnosed case in between them in the transmission chain to be classified as a single household cluster.

Variables collected during routine surveillance and utilized in this analysis were age, gender, race/ethnicity, address, symptom status, hospitalization, date of positive sample collection and date of symptom-onset. We estimated the extent of COVID-19 clustering among cases by calculating the proportion of all COVID-19 cases that belong to a household cluster, based on our operational definition of a cluster described above, stratified by age group, gender, race/ethnicity, symptom and hospitalization status and over month of the pandemic. We computed statistics related to cluster characteristics including cluster size and duration between first and last diagnosed case within a cluster. We focus our analysis on patterns in age profiles of cases in household clusters by describing the distribution of the age of first diagnosed case within a household cluster over time and visualizing patterns between age group of first diagnosed case and age group of subsequent household cases. For the latter visualization, we expected age-specific relationships between first and subsequent cases within households to mirror the age-specific mixing patterns within household contacts documented in social mixing surveys^25^.

This activity was determined to be consistent with public health surveillance activity as per title 45 code of Federal Regulations 46.102(l)(2). The Emory University institutional review board approved this activity with a waiver of informed consent.

## Results

A total of 106,233 COVID-19 cases were reported in Fulton County between 6/1/20-10/31/21, of which 84,383 (79.4%) were included in our analysis. Reasons for exclusion include residing in a long-term care facility (N=4,540), addresses not matched to the geocoding database (N= 5,991), live in communal places (ex. correctional facilities, shelters or dorms) (N= 1,509) and apartments with no unit number (N= 9, 810) (Supplemental Figure 2). Included individuals were similar to excluded individuals with respect to age, gender, race/ethnicity, symptom and hospitalization status (Supplemental Table 1). A majority of cases were female (N= 44,940; 53%), 14,726 (17%) were children aged 0-18 years and 10,555 (12%) were adults aged 60 years and above (Table 1). The largest racial/ethnicity group was black, non-Hispanic individuals (N = 38,128, 45%), followed by white, non-Hispanic individuals (N=28,272, 34%) followed by Hispanic individuals of all races (N= 7776, 9%).

**Table 1.** Demographic characteristics of confirmed COVID-19 cases with a valid residential address and proportion of cases identified in household clusters – June 1, 2020 to October 31, 2021

|  |  | Total | Col% | HH-clustered<br>individuals<br>(n=27,898) | % HH-clustered |
| --- | --- | --- | --- | --- | --- |
| **Total population** |  | **84,383** | **--** | **31,449** | **37** |
| **Gender** | Female | 44940 | 53 | 16944 | 38 |
|  | Male | 39150 | 46 | 14408 | 37 |
|  | Missing | 293 | 0 | 97 | 33 |
| **Age group** | 0-18 | 14726 | 17 | 8089 | 55 |
|  | 19-29 | 18647 | 22 | 5080 | 27 |
|  | 30-39 | 16125 | 19 | 4820 | 30 |
|  | 40-49 | 13050 | 15 | 5021 | 38 |
|  | 50-59 | 11254 | 13 | 4298 | 38 |
|  | 60-69 | 6263 | 7 | 2418 | 39 |
|  | 70 plus | 4292 | 5 | 1715 | 40 |
| **Race/ethnicity** | Asian, NH | 3503 | 4 | 1693 | 48 |
|  | Black, NH | 38128 | 45 | 13339 | 35 |
|  | White, NH | 28272 | 34 | 10038 | 36 |
|  | Hispanic, all | 7776 | 9 | 3769 | 48 |
|  | Other, NH | 2678 | 3 | 1073 | 40 |
|  | Missing | 4026 | 5 | 1537 | 38 |
| **Symptom status** | No | 8812 | 10 | 3299 | 37 |
|  | Yes | 49247 | 58 | 18468 | 38 |
|  | Unknown | 26324 | 31 | 9682 | 37 |
| **Hospitalized** | No | 46602 | 91 | 17974 | 39 |
|  | Yes | 4758 | 9 | 1476 | 31 |

Among these cases, 37,432 (44%) had an address that matched at least one other case; 31,449 (37%) (**Table 1**) had positive sample collection date within 28 days of another household case. The age-stratified probability of being part of a household cluster followed a U-shaped trend where children aged 0-18 years were most likely to be part of a household cluster (55%), followed by adults aged 40 years and above (39%), with young adults between 19-39 years the least likely to be in a household cluster (28%). We observed higher clustering among Hispanic (47%) and non-Hispanic, Asian (47%) persons compared to non-Hispanic black (33%) and non-Hispanic white (35%) persons. There are no differences in clustering by gender or reported symptom status.

We observed temporal trends in household clustering. The proportion of household cases identified in clusters fluctuated between 30-40% each month. Clustering increased between November 2020 to January 2021, steadily declined between Feb to June 2021, arrived at a low in June 2021, and has since rebounded to earlier levels (**Fig 1**). Trends in probability of household clustering by age of case (ex. children aged 0-18 years most likely to be in household clusters) have stayed consistent over the course of the pandemic (**Figure 2**).

**Figure 1.**
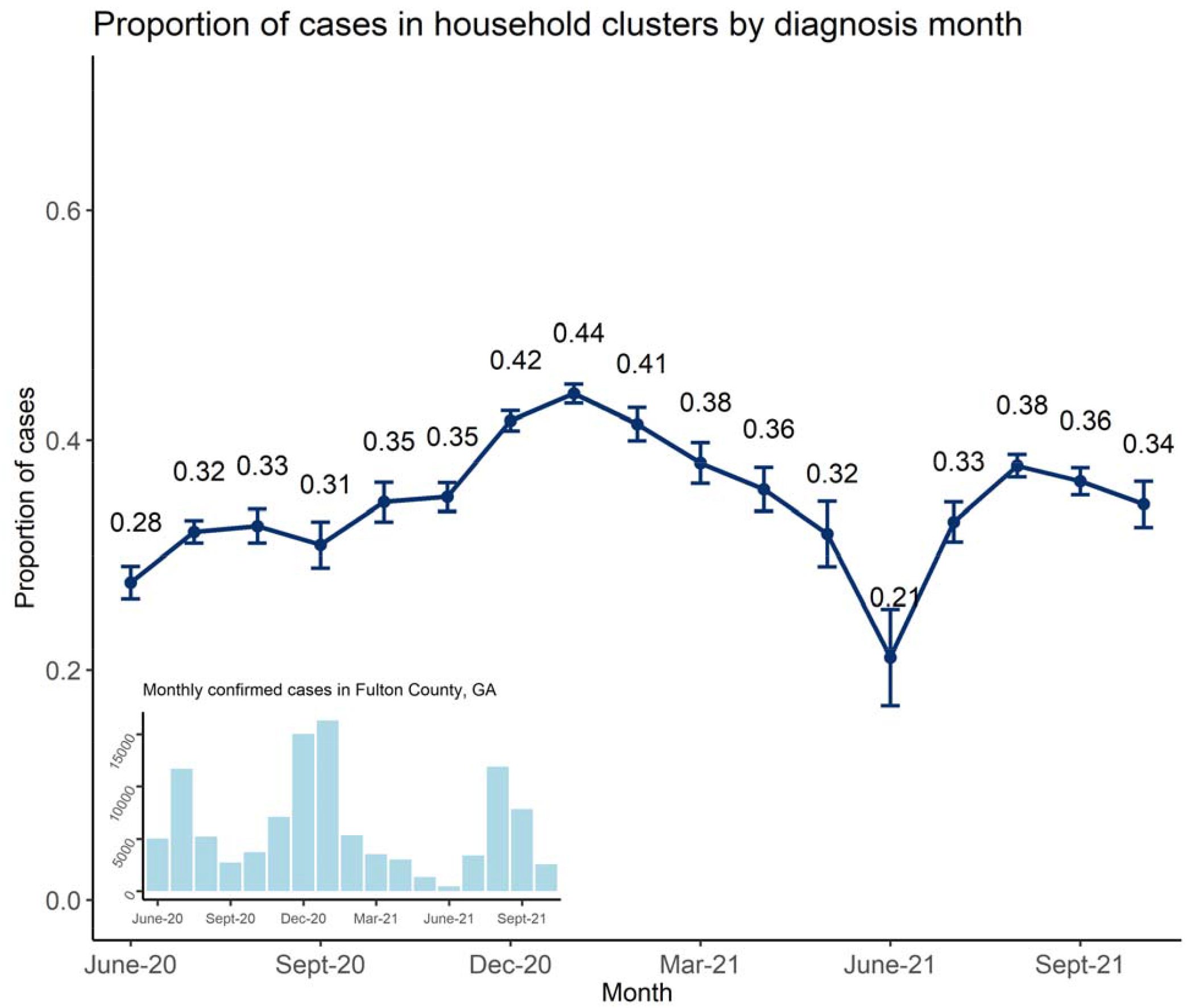
Temporal trend in the proportion of diagnosed cases in Fulton County, Georgia, that were identified in household clusters stratified by the month of positive sample collection date (dark blue line), with 95% confidence interval around the point estimate. A bar chart of monthly confirmed cases in Fulton County is provided on the bottom left for reference.

**Figure 2.**
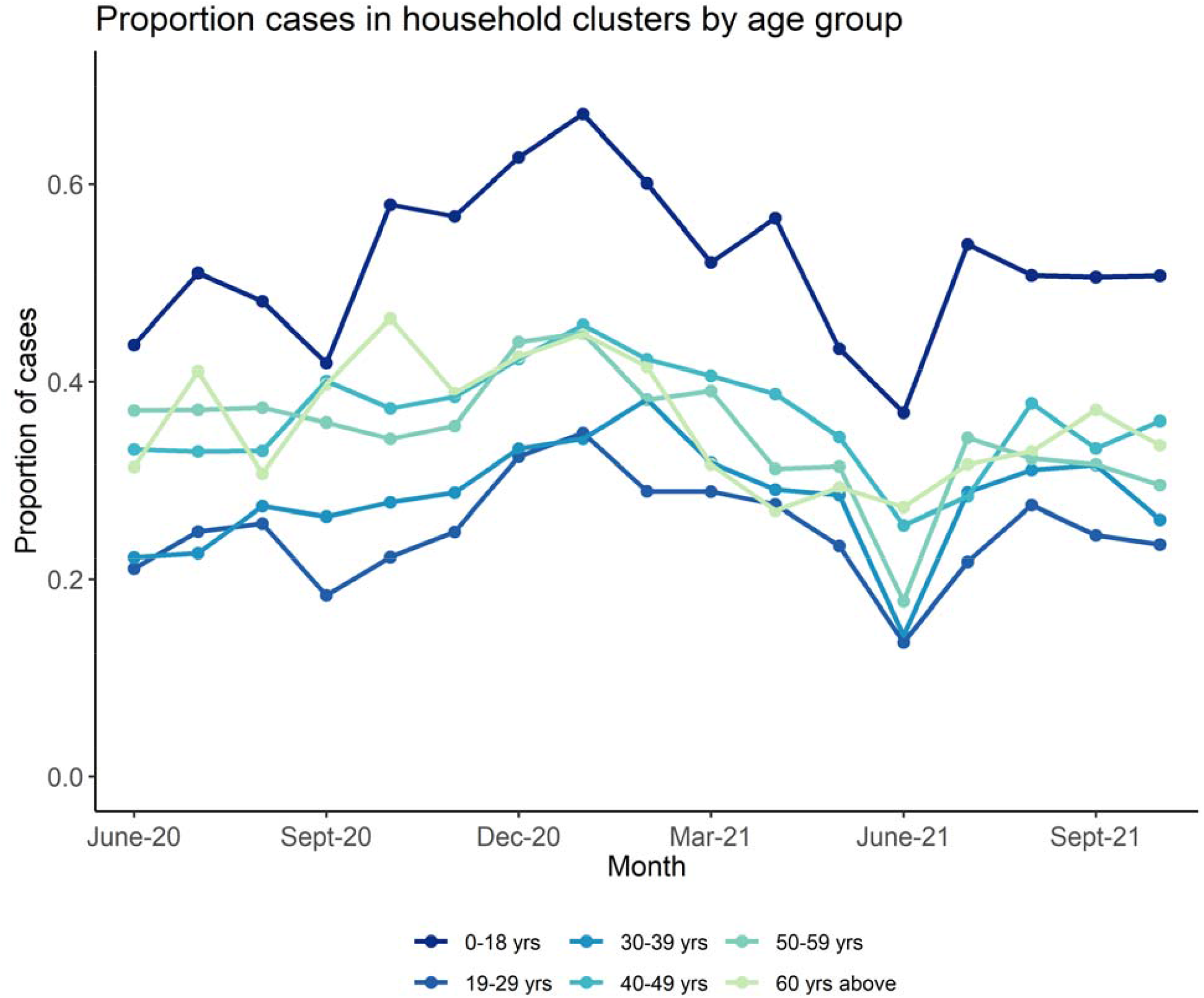
Temporal trend in the proportion of diagnosed cases in Fulton County, Georgia, that were identified in household clusters stratified by month of positive sample collection date (x-axis) and by age group

Among 12,955 household clusters, the majority of clusters had 2 individuals (N=9216 clusters, 71%), although some clusters had ≥6 individuals (N=122, 1.0%; **Table 2**). Excluding clusters with multiple cases diagnosed on the first day, the first diagnosed case was 0-18 years in 1,314 (15%) of clusters, 19-29 years in 1,614 (19%) of clusters, 30-39 years in 1,592 (18%) of clusters, 40-49 years in 1,593 (18%) of clusters 50-59 years in 1,336 (15%) of clusters and above 60 years in 1,235 (14%) of clusters. The proportion of children diagnosed as the first case in the cluster increased from 11% in February 2021 to a high of 31% in August 2021 (**Figure 3**). In contrast, proportion of elderly greater than 50 years of age diagnosed as the first case in the cluster decreased during the same time period from 34% to 19%. Clusters most often consisted of individuals in the same age group, as shown by the high density of bubbles along the diagonal in the bubble plot counting clusters by age of first diagnosed case and age of subsequent diagnosed cases in the household (**Figure 4**). Two other diagonals are present. One below the main diagonal consisting of 25-50-year-old first diagnosed case clustered with 0-20-year-old subsequent household cases and another diagonal above the main diagonal consisting of 10-25-year-old first diagnosed case clustered with 40-55-year-old secondary cases.

**Table 2.**
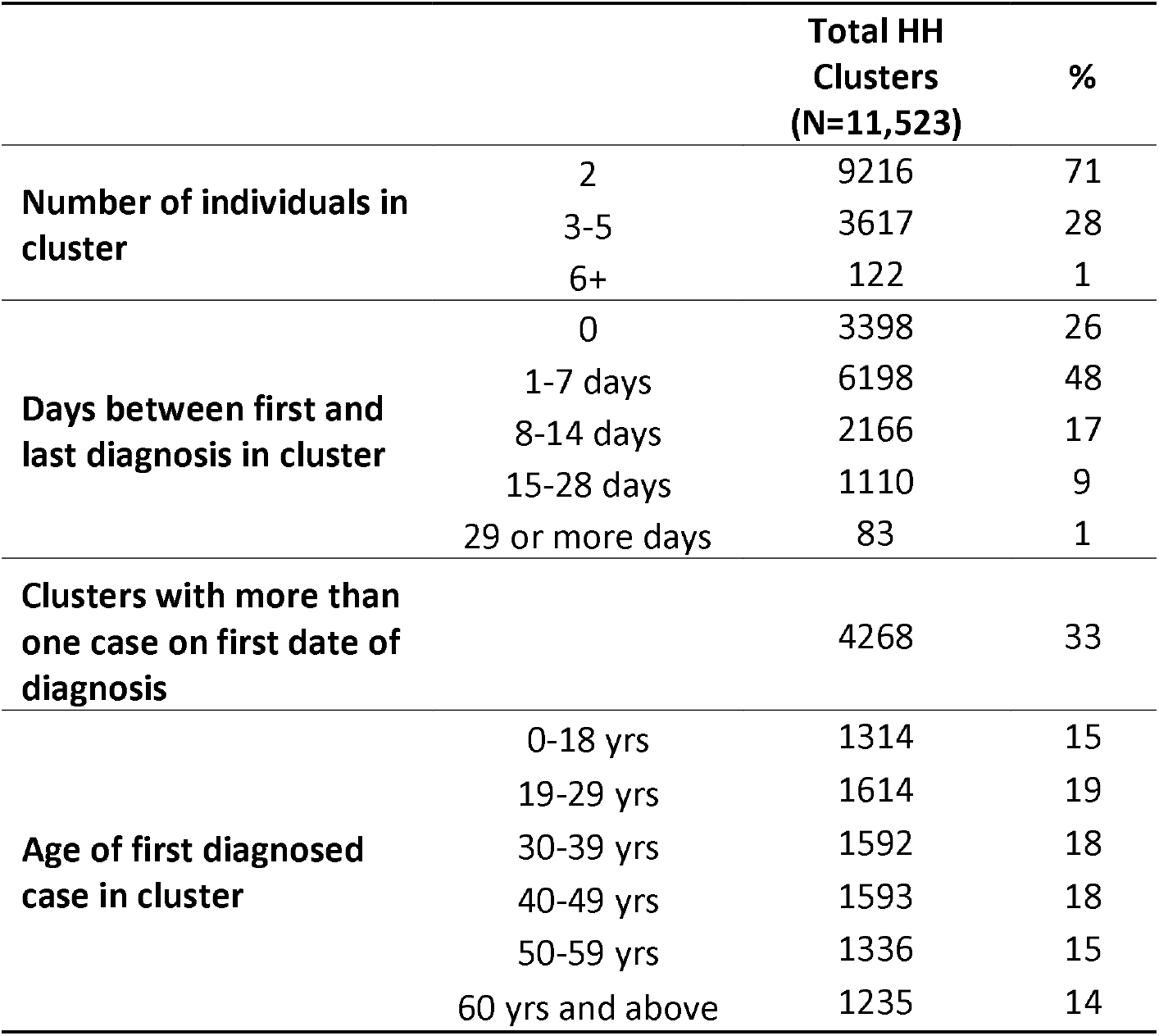
Characteristics of household clusters (N= 11,523) in Fulton County – June 1, 2020 October 31, 2021.

**Figure 3.**
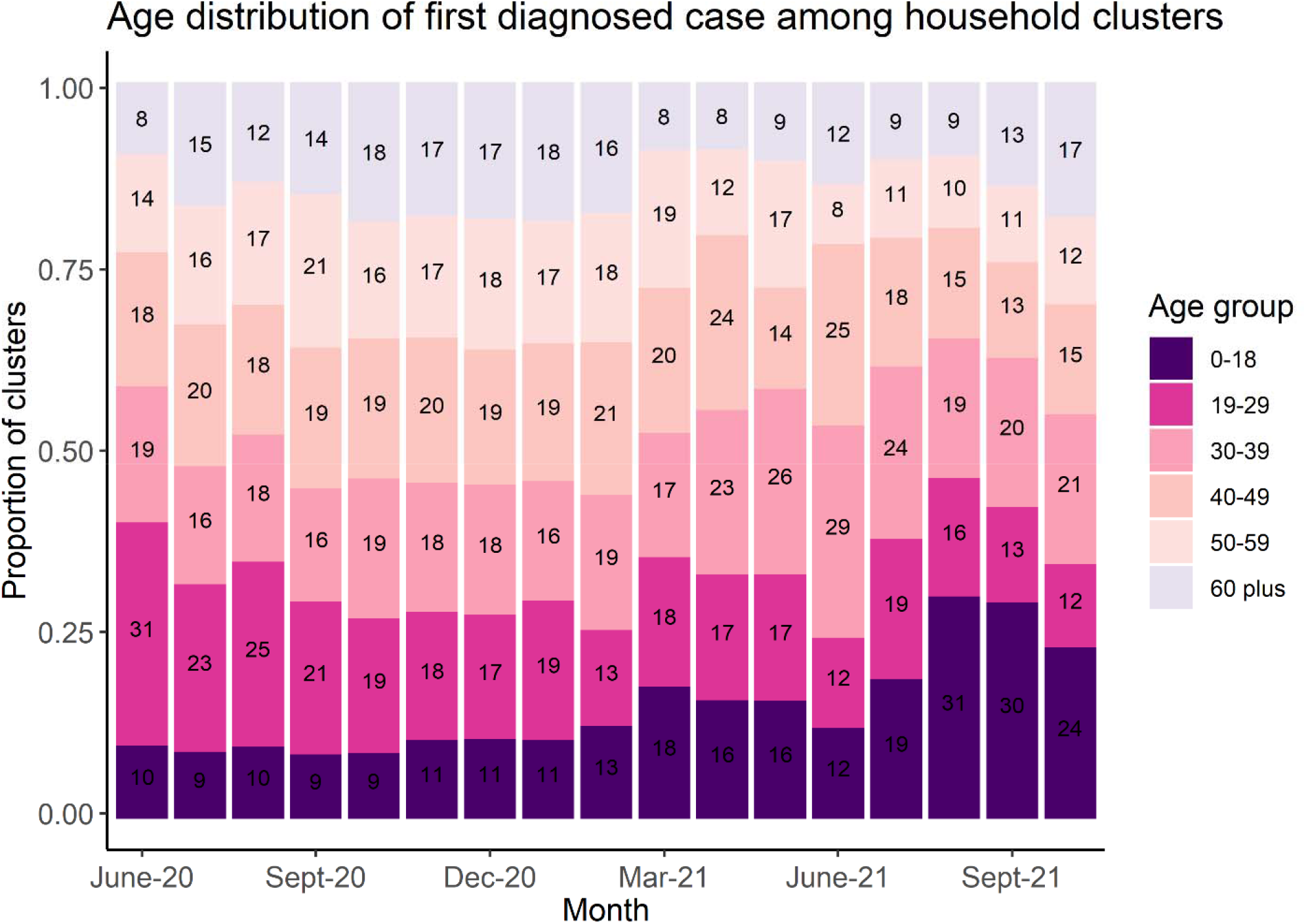
Temporal trend in the age distribution of first diagnosed case among household clusters (N= 7689) identified in Fulton County, Georgia between June 1, 2020 and Oct 31, 2021

**Figure 4.**
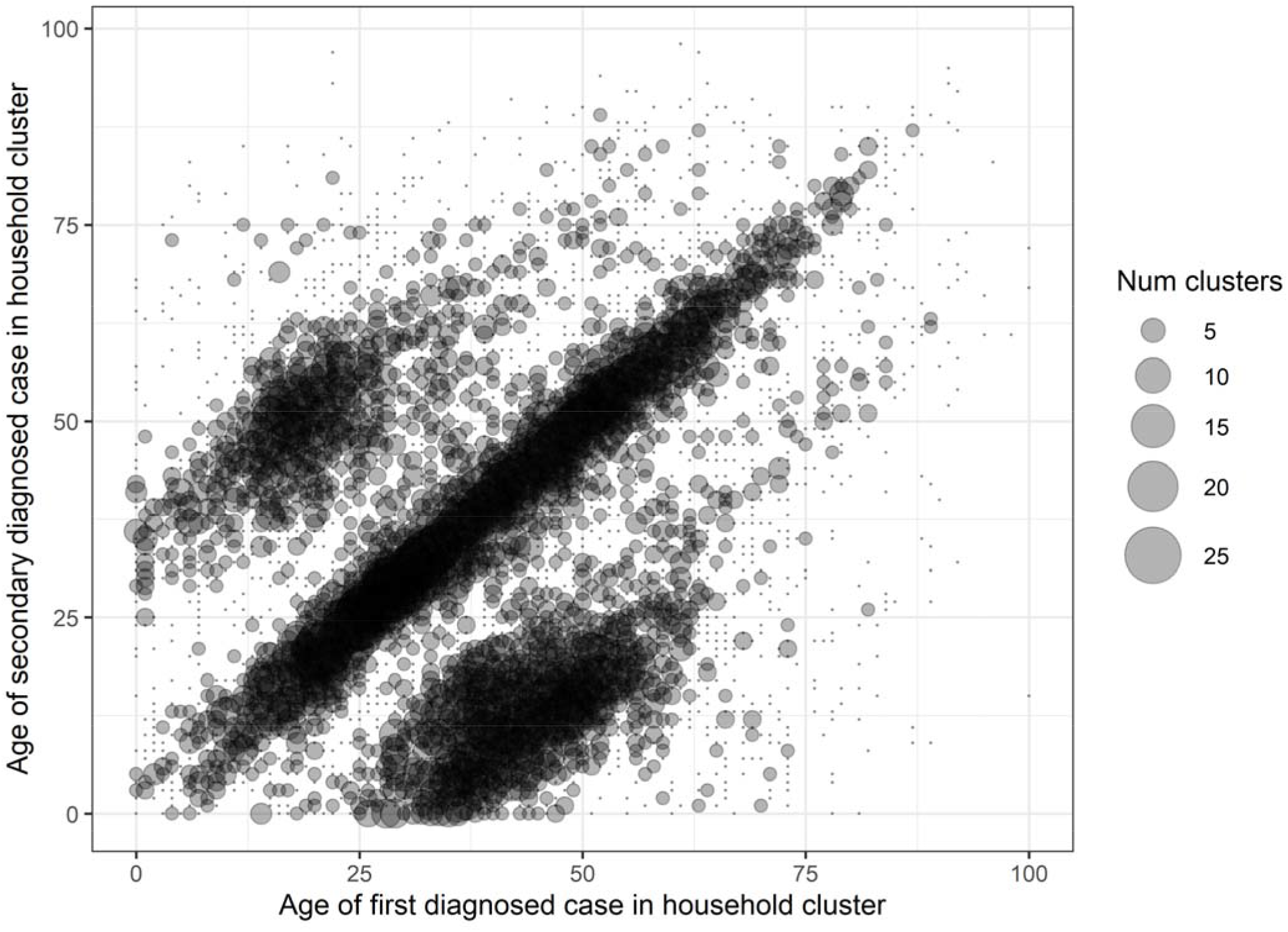
Bubble plot of distribution of clusters across different cluster age profiles where the x-axis is the age of first diagnosed case in the household cluster, y-axis is the age of subsequent secondary diagnosis in the household cluster, the size of bubble represents number of clusters by each age pairing of first diagnosed case and subsequent cases and the density representing more common cluster age profiles (on the diagonal between cases of the same age in the same household and the two off-diagonal “wings” representing intergenerational clusters).

## Discussion

Over one-third of reported COVID-19 cases in Fulton County between June 2020 and August 2021 were part of a household cluster. Children aged 0-18 years were substantially more likely to belong to a household cluster than any other age group and increasingly represented the first positive detected case in the household, rising from 11% in February 2021 to 31% in August 2021. Age patterns between first and subsequent cases within household clusters mirror household social mixing patterns where clusters of individuals of the same age are most common followed by intergenerational clusters between parents and children. High probability of household clustering and cross-age transmission underscore the importance of contact tracing, testing, and quarantining of household contacts and the need for innovative strategies such as self-testing of household members for early identification of infections.

Findings from our study adds evidence to the outsized role of household clustering in shaping the course of the pandemic. Our study results are comparable to those reported by Massachusetts State Department of Public Health in their “cluster-busting” strategy for COVID-19 control^34^ where a third of cases reported between September and October 2020 belonged to a household cluster^35^. Small reductions in household transmission have potential to meaningfully reduce overall cases. On a daily basis, a larger proportion of close proximity human contact occurs within households^36–38^ rather than in community settings such as schools, nursing homes or large gatherings notorious for superspreading. While households rarely become superspreading locations, the majority of Americans (72%)^39^ live with at least one other individual who would be highly exposed to an index household case.

The higher proportion of children in household clusters likely reflects higher probability of living in a home with another individual, either other children or adult caregivers. Increased probability of child cases as the first diagnosed case in household clusters starting March 2021 coincide with vaccination of older age groups. Further increases starting in August 2021 coincide with the return-to-school of largely unvaccinated children for the Fall 2021 semester. Collectively, these trends suggest that children are increasingly important for transmission within households despite lower infectiousness compared to adults.^40,41^ Our bubble plot of age profiles within household clusters show that clusters are dominated by those of individuals of the same age, likely couples, roommates or sibling, and those in intergenerational age groups, likely parent and child. These findings suggest the importance of age-specific mixing patterns in determining the magnitude of age-specific clustering within households.

As the COVID-19 pandemic continues, household-level interventions to reduce household clustering should be further incorporated into the existing response. For example, the US Centers for Disease Control and Prevention recommends household index cases quarantine in a separate bedroom and bathroom and limit sharing of food and kitchenware.^42^ Yet distancing and safe quarantine are not feasible within all households and an estimated 20% of households did not have sufficient bedrooms and bathrooms to safely quarantine an infected person at home.^43^ Vaccination remains the single most effective intervention at preventing infection and severe disease;^44^ however, while coverage among children is still low,^45^ additional interventions should be explored such as rapid antigen tests for all household members to facilitate early diagnosis or provision of a box of surgical or KN95 masks to increase safer household contact and prevent onward transmission outside of the household.

We report several limitations. Our analysis used surveillance data which is known to under-ascertain COVID-19 cases, especially during the early days of the pandemic^46^. Individuals with known household exposure may be more likely to present for testing if health conscious or less likely to present if they believe knowing their infection status will have little impact on treatment course and outcome. Changing testing and screening strategies may also affect age-related clustering trends. Moreover, Children 0-18 years were slightly more likely to be included compared to young adults aged 19-29 year and the elderly aged 70 years and above. Furthermore, we do not know if members within household clusters infected each other or whether some subsequent cases were infected from the community. The distribution of infections caused by household exposure versus community exposure has implications for disease control strategies. Finally, the first diagnosed case may not represent the source of infection for the household.

The unique advantage of our study is that we use rigorous methods to identify cases from surveillance data residing at the same residential address, producing the first estimates of the extent of household clustering over time in a large, diverse metropolitan area. No other study has used public health surveillance data to systematically track temporal and demographic trends of household clusters of COVID-19 in the US. The use of routinely collected surveillance data provides a more accessible, rapid approach for health departments to evaluate household clustering and inform interventions. In addition, our study finds evidence of higher probability of household clustering among children and Hispanic and Asian persons and early indication of increases in the proportion of household clusters with a child as the first diagnosed case.

In conclusion, we used residential address to identify cases temporally clustered within a household. Our analysis found that between June 1, 2020 and October 31, 2021, 37% of reported cases in Fulton County, Georgia belonged to household clusters. Our findings complement the high household secondary attack rates found in cohort studies of household transmission of SARS-CoV-2 and further quantifies the extent of household clustering at a population level. Our results support the consideration of improved public health response to reducing household clustering and within-household transmission such as within-household masking and rapid testing for household contacts for far-reaching public health impact in controlling COVID-19.

## Data Availability

Data produced in the present study are available upon reasonable request to the authors.

## Supplementary Information

**Supplemental Figure 1.**
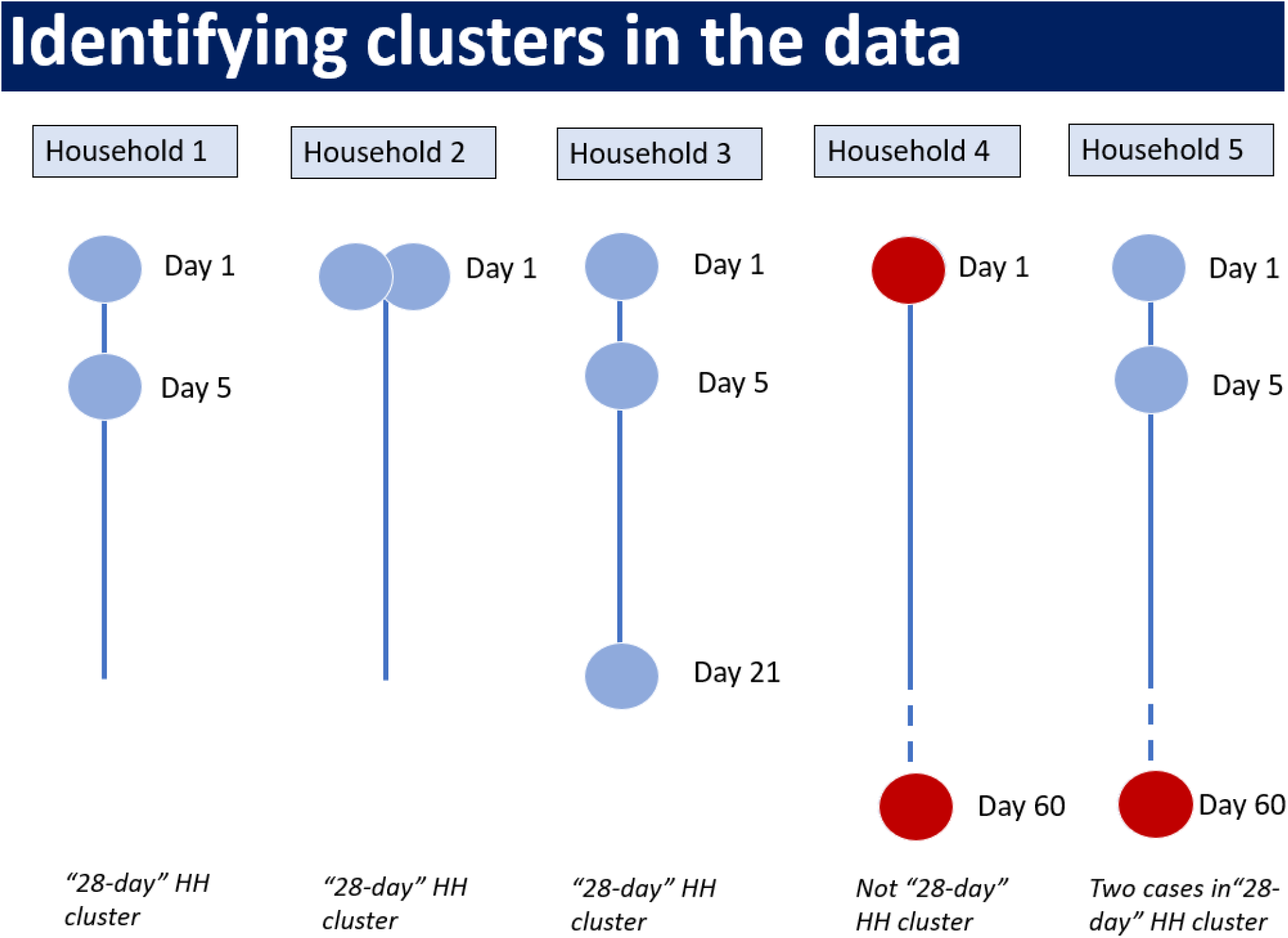
Identifying clusters in the dataset and illustrations of scenarios that we consider as a household cluster versus scenarios that we do not consider as a household cluster. Our operational definition of household-clustered COVID-19 cases is two or more cases reporting a valid address at the same residential location less than 28 days apart. The days are by date of positive sample collection and day 1 is the date of positive sample collection of first identified case at address.

**Supplemental Figure 2.**
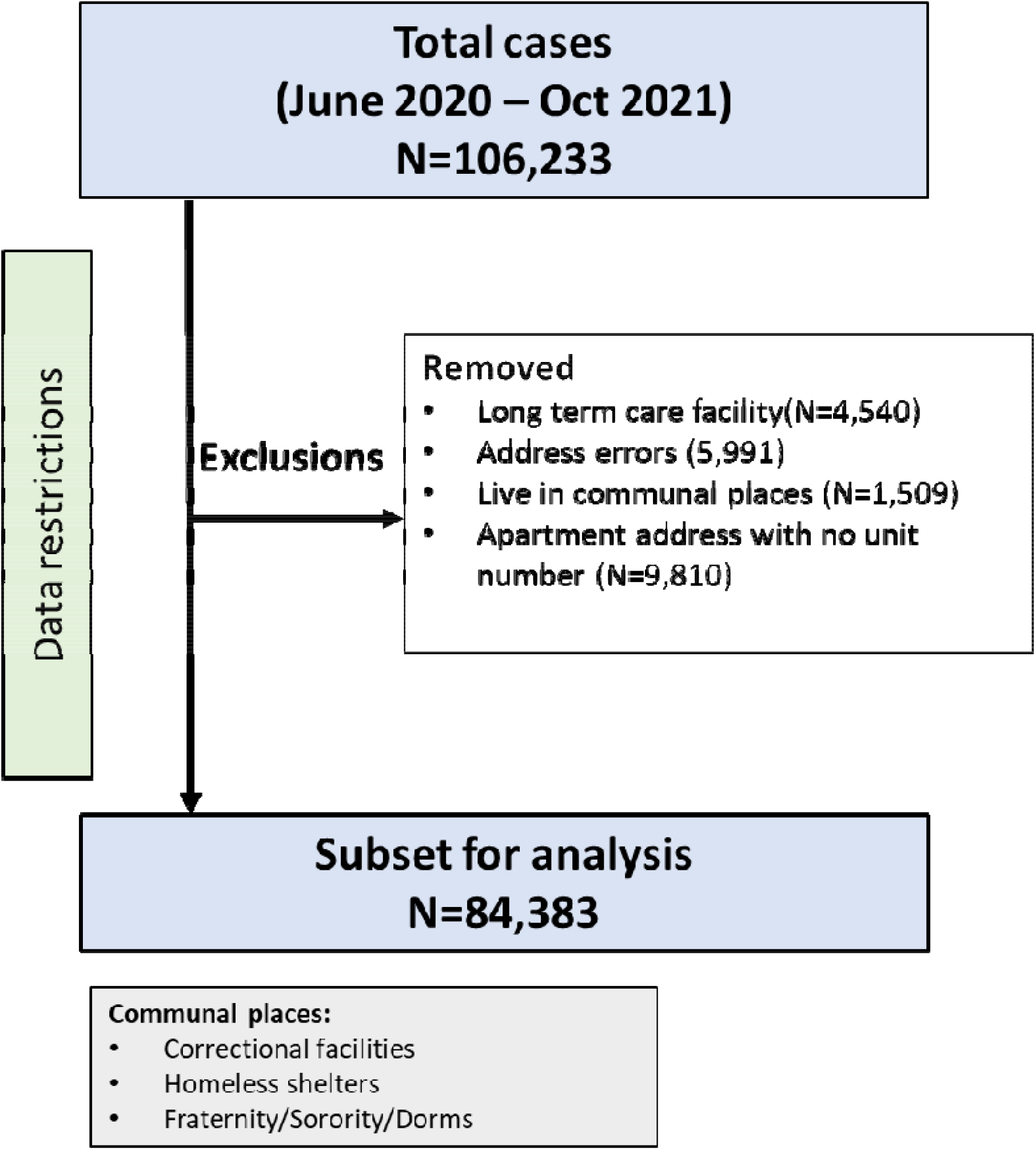
Flow diagram of cases included in the study

**Supplemental Table 1.**
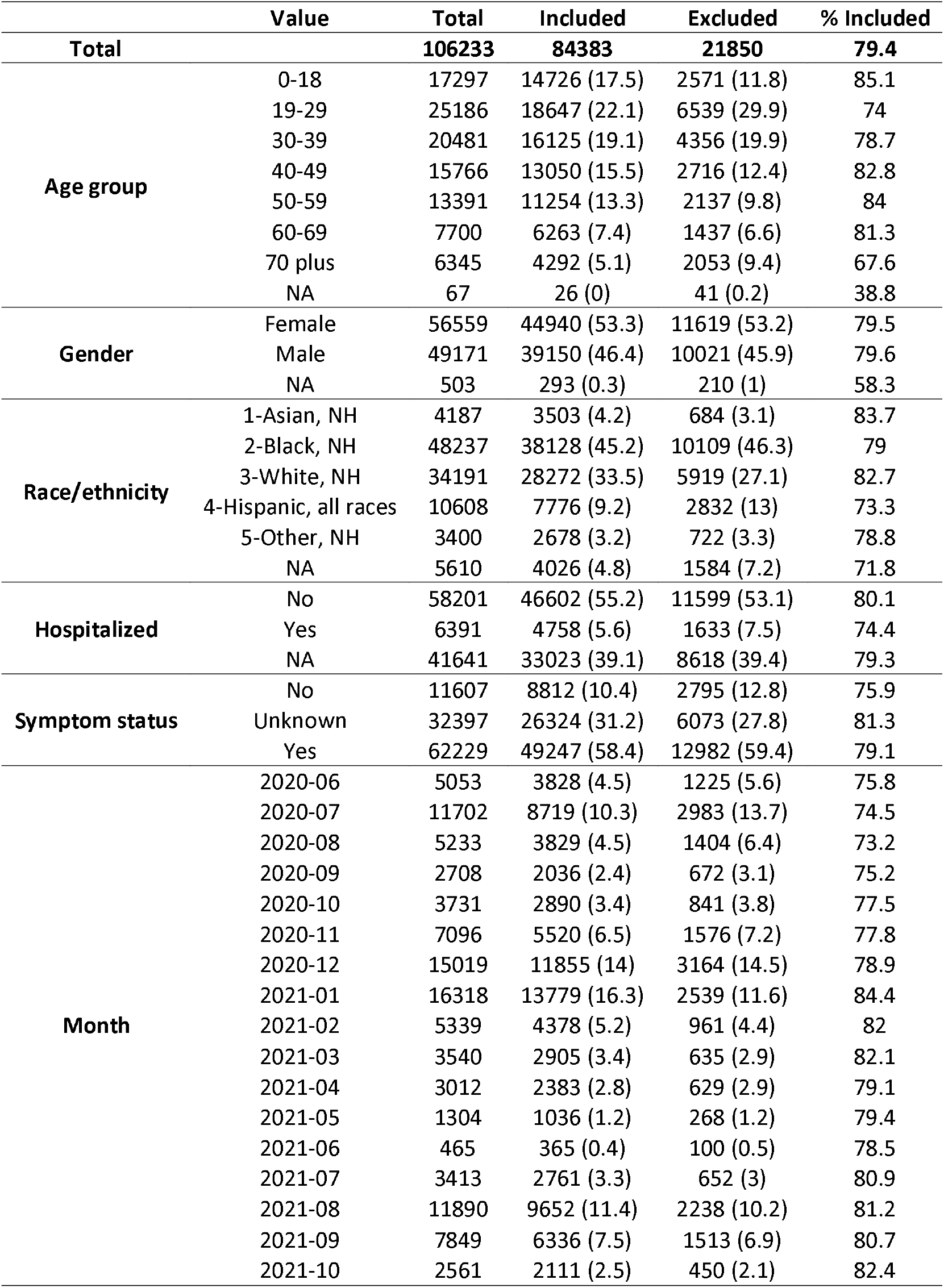

**Supplemental Figure 3.**
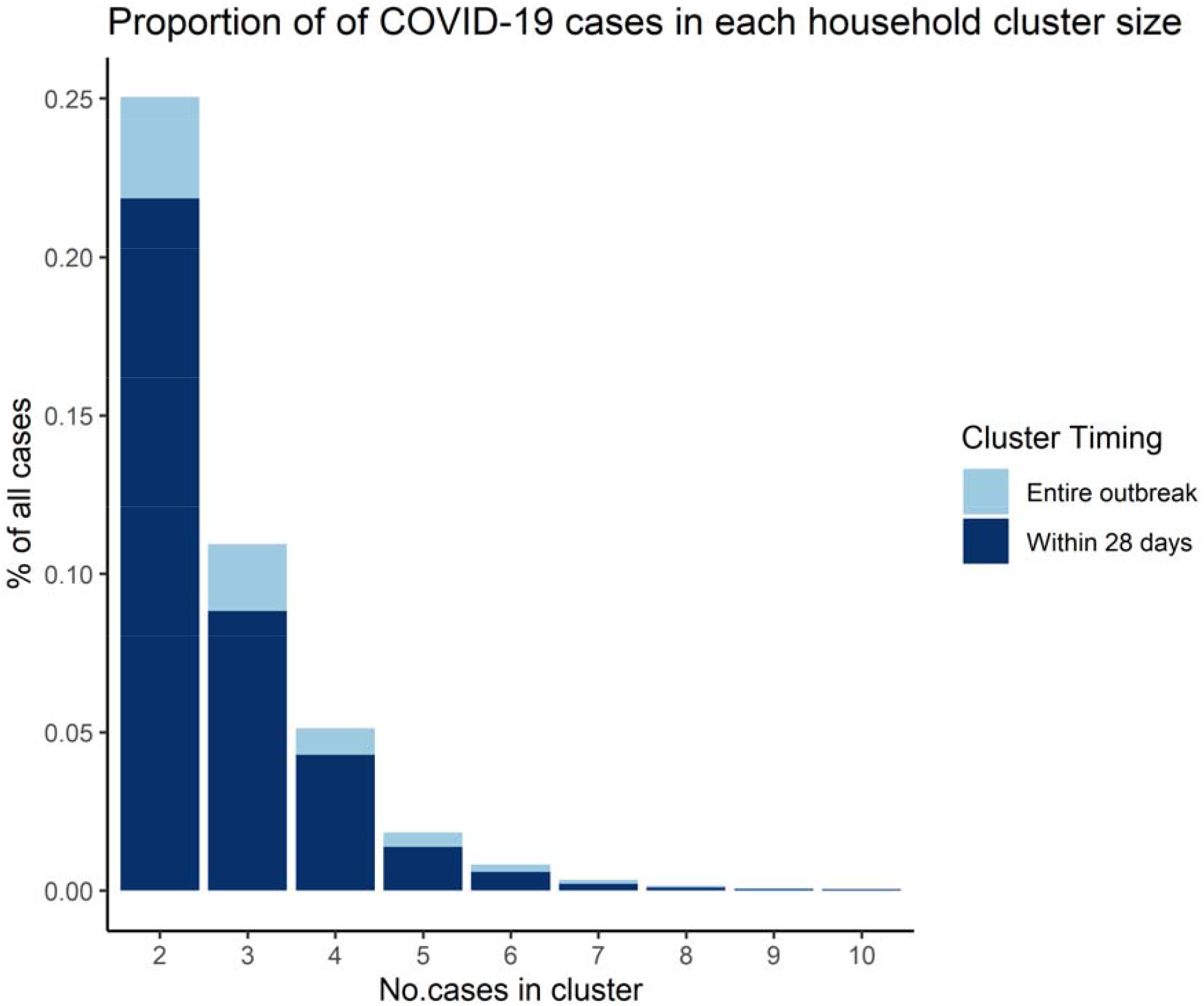
Proportion of COVID-19 cases in each household cluster size when clusters were counted for the entire outbreak versus when clusters were restricted to those within 28 days of subsequent positive sample collections

**Supplemental Figure 4.**
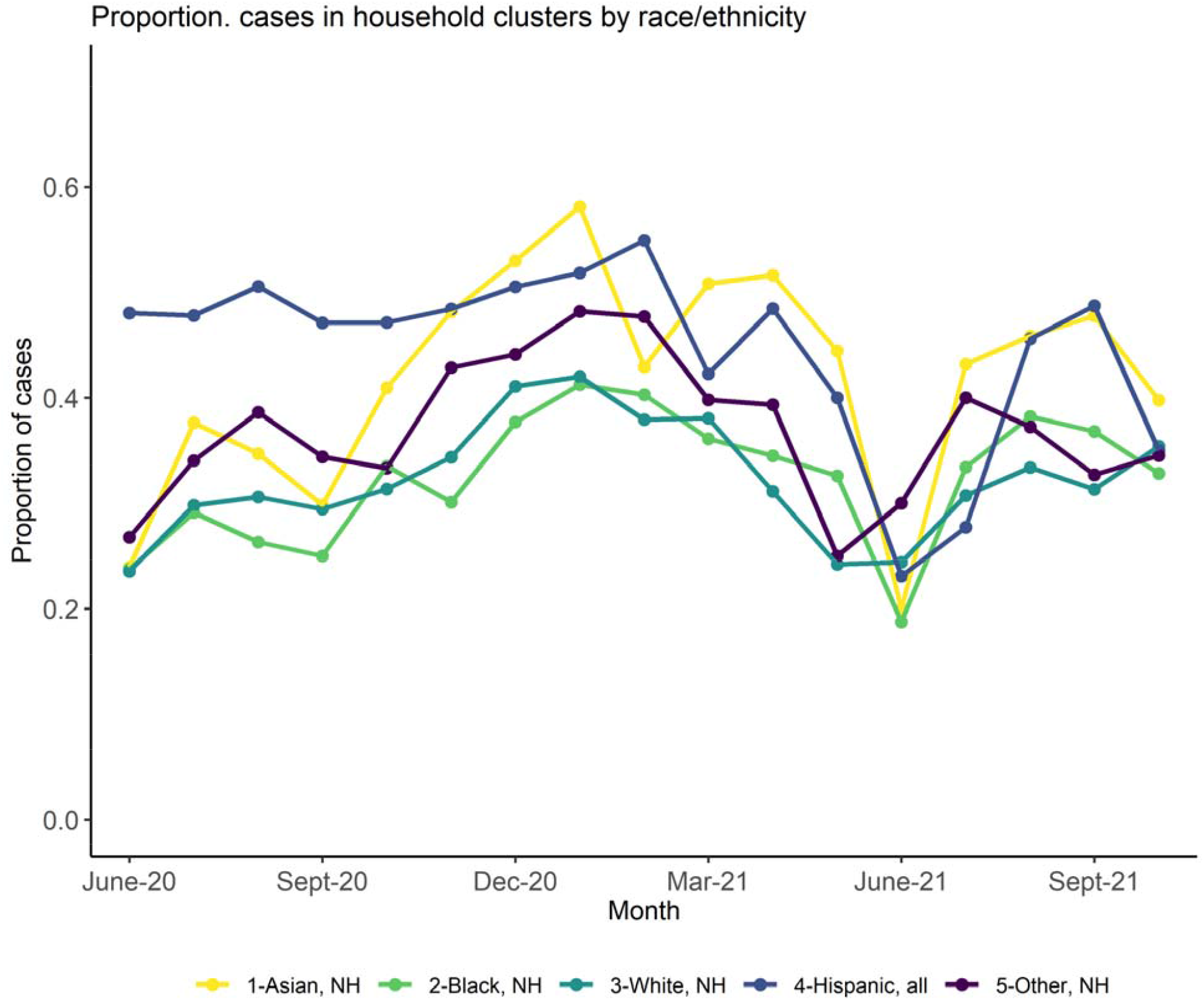
Temporal trend in the proportion of diagnosed cases in Fulton County, Georgia, that was identified in household clusters stratified by month of positive sample collection date (x-axis) and race/ethnicity.

**Supplemental Table 2.** Average number of household members (all ages and children 0-18 years) per household by race/ethnicity.

|  | Average number household<br>members (all ages) | Average number of children<br>(0-18 years) per household |
| --- | --- | --- |
| All races | 2.36 | 0.58 |
| Asian | 2.79 | 0.74 |
| Black | 2.41 | 0.65 |
| Hispanic | 3.38 | 1.11 |
| White | 2.22 | 0.47 |

